# Patterns in Patient Portal Enrollment and Use Among Hospitalized Children

**DOI:** 10.64898/2026.01.09.26343716

**Authors:** Michael J. Luke, Yinlu Zhu, Courtney B. Wolk, Stephanie G. Menko, Danielle Capriola, Katherine Fuller, Philip V. Scribano, Christopher P. Bonafide, Aditi Vasan

## Abstract

**Background and Objectives:** Patient portals allow adolescents and caregivers of pediatric patients to better engage with health care, improving their medication adherence, health knowledge, self-efficacy, and receipt of preventive care. However, disparities in access to and comfort with using these portals persist. Hospitalization represents a promising opportunity to address these disparities among a high-risk population.

**Design/Methods:** We conducted a cross-sectional analysis to identify disparities in portal enrollment and use (logins, messaging, or telehealth use) among patients admitted to two pediatric hospitals within a single health system from 2022-2024. We calculated unadjusted rates of portal enrollment and use before, during, and after hospitalization, stratified by patient-level factors (age, race/ethnicity, insurance coverage, medical complexity), household-level factors (preferred language), and population-level factors (neighborhood opportunity). We then used multivariable logistic regression to identify associations between these factors and portal enrollment and use.

**Results:** Among 40,371 hospitalized patients, 93% had enrolled in our patient portal. Patients who identified as Non-Hispanic Black, were publicly insured, had a household language other than English, and lived in lower opportunity neighborhoods had significantly decreased odds of portal enrollment and use before, during, and after hospitalization.

**Conclusions:** Despite high overall rates of portal enrollment among our patient population, we observed persistent disparities in portal enrollment and use. Efforts to promote equitable portal usage among hospitalized children may be most effective when focused on families who are publicly insured, prefer languages other than English, or live in lower opportunity neighborhoods.

## Introduction

Digital patient portals allow caregivers of pediatric patients to engage with their children’s health care via a growing range of features, including telehealth visits, bi-directional messaging with providers, and the ability to schedule appointments and request medication refills. Patient portals also facilitate easy access to medical records, laboratory results, and health education resources.^1, 2^ Patient portal use has been associated with improvements in medication adherence, health knowledge, self-efficacy, and engagement with preventive care.^3-5^ However, not all eligible families activate or engage with their child’s patient portal, and some families may face structural barriers. Several prior studies have described disparities in patient portal enrollment and use among adults, with lower rates of patient portal use among low-income and minoritized individuals.^6-8^

Patient portal use may be particularly beneficial for children and their caregivers, given the frequent preventive services and screenings needed in childhood.^9^ Patient portal use may also be especially important during pediatric hospitalizations, allowing caregivers to monitor their child’s progress in real-time and make more informed decisions.^10^ Some recent studies have examined disparities in patient portal access in inpatient pediatric settings;^11^ however, these studies have been limited in the diversity of their patient populations and have focused more on portal enrollment than on meaningful use.^12, 13^ We therefore aimed to characterize disparities in patient portal enrollment and use among families of hospitalized children at two pediatric hospitals within a single health system. We plan to use insights from this descriptive study to inform future targeted quality improvement efforts aimed at equitably improving patient portal engagement and mitigating disparities.

## Methods

We conducted a cross-sectional observational study exploring the association of patient-level, household-level, and population-level factors with pediatric patient portal enrollment and usage before, during, and after hospitalization.

### Setting and Participants

We included all patients who were admitted to an urban quaternary care children’s hospital or an affiliated suburban tertiary care children’s hospital from 2022 to 2024 who had been seen by either a primary care pediatrician or at least one subspecialist affiliated with our health system. We excluded patients without a primary care pediatrician or subspecialist affiliated with our health system as they might receive most of their care through another health system and may therefore receive fewer benefits from our patient portal.

We included only the first admission for each patient during our study period and excluded patients without at least 3 months of post-discharge portal data available. Our health system uses the Epic Electronic Health Record system and patient portal, which allows caregivers to access their children’s portals as proxies until the age of 13, after which adolescents have private access to the portal.^14^ We included both caregiver proxy access and adolescent patient access in our study. Patients and caregivers can be enrolled by request or recommendation from their providers during clinical encounters, by front-desk staff in our primary care or subspecialty clinics, or by registration staff in our emergency departments or hospital units. During this study period, there was no mandated or consistent process for patient portal enrollment across our health system.

### Measures

Our primary outcomes were patient portal enrollment prior to hospitalization, patient portal enrollment during hospitalization, and patient portal usage assessed at 3 time points:

12 months prior to admission, during the hospitalization, and 3 months after the hospitalization. Patient portal usage was defined based on whether the patient or their caregiver proxy had one or more patient portal logins, sent messages, or attended telehealth visits during the specified time period.

We examined several patient-level factors that we hypothesized might be associated with patient portal enrollment and use, including patient age (dichotomized as < 13 or >= 13), race/ethnicity, insurance coverage (public vs. private), and medical complexity, measured using a binary indicator for whether each child had been diagnosed with one or more Complex Chronic Conditions (CCC) classified using CCC algorithm version 3.0.^15^ Age was dichotomized as <13 or >=13 in line with age categories for proxy and adolescent portal access. However, for patients >=13, our data do not distinguish between use by patient or proxy. As a household-level factor associated with patient portal enrollment, we examined the household’s preferred language for receiving medical care, operationalized as a binary variable with categories of English or any language other than English (LOE). Lastly, as a population-level factor associated with patient portal enrollment, we examined ^15^neighborhood opportunity at the census tract level, approximated using nationally-normed Child Opportunity Index (COI) 3.0 quintiles based on residential address on file.^16^ Race/ethnicity and COI were used as imperfect proxy measures for structural racism and discrimination, which may influence both families’ likelihood of being referred for patient portal enrollment and their likelihood of engaging with the patient portal via mechanisms such as trust and health literacy.

### Analysis

We used descriptive statistics to characterize our population and describe how rates of patient portal enrollment and use varied based on the patient-level, household-level, and population-level factors described above. We then used multivariable logistic regression to examine the association of these patient-level, household-level, and population-level factors with portal enrollment and use. In all multivariable regression models, we included fixed effects for hospital site and calendar year, to account for site-based differences and trends in patient portal enrollment and use over time. In addition, in our multivariable models examining patient portal enrollment and usage during hospitalizations, we adjusted for length of stay in quartiles, as we hypothesized that caregivers of children with longer hospitalizations may be more likely to be enrolled in and utilize the patient portal while hospitalized. In our multivariable analysis, we excluded race or ethnicity groups representing <1% of our study sample. Consistent with best practices, we used a composite categorical variable for race and ethnicity and assigned the group with greatest adjusted odds of our primary outcome (portal enrollment) as our reference category (patients who identified as Asian).^17, 18^ Similarly, for COI, we used the highest quintile (“Very High”) as our reference. All analyses were performed using Stata 18.0.^19^

## Results

Our study sample included a total of 40,371 unique patients (Table 1). Overall, 47% identified as Non-Hispanic White, 44% were publicly insured, 5% preferred a language other than English, and 46% had one or more CCCs, representative of the increased prevalence of CCCs in the past decade.^20^ The vast majority of our sample was enrolled in the patient portal prior to their child’s admission (93%), but only 40% had sent one or more messages, and 17% had used telehealth in the previous year (Table 2).

**Table 1:** Sample Characteristics.

| **Patient-, Household-, and Population-Level Factors** |  | **Overall** |
| --- | --- | --- |
|  |  | **N = 40,371** |
| **Age Category** | <13 (Caregiver Access) | 30,147 (75%) |
|  | >= 13 (Patient and Caregiver Access) | 10,224 (25%) |
| **Race and Ethnicity** | Asian | 1,772 (4%) |
|  | Non-Hispanic White | 18,813 (47%) |
|  | Hispanic or Latino | 5,222 (13%) |
|  | Non-Hispanic Black | 10,068 (25%) |
|  | American Indian or Alaskan Native | 24 (0%) |
|  | Native Hawaiian or Pacific Islander | 13 (0%) |
|  | Multi-Racial | 1,717 (4%) |
|  | Other | 2,257 (6%) |
|  | Unknown | 485 (1%) |
| **Insurance Status** | Public | 17,757 (44%) |
|  | Private | 22,565 (56%) |
|  | Other | 49 (0%) |
| **Complex Chronic Conditions** | 1+ | 18,547 (46%) |
| **Preferred Language** | English | 38,165 (95%) |
|  | Spanish | 1,304 (3%) |
|  | Other Non-English Language | 902 (2%) |
| **Child Opportunity Index** | Very Low | 7,638 (19%) |
|  | Low | 4,616 (11%) |
|  | Moderate | 4,706 (12%) |
|  | High | 7,793 (19%) |
|  | Very High | 15,618 (39%) |
| **Hospital Site** | Suburban Community Site | 7,783 (19%) |
|  | Urban Academic Site | 32,588 (81%) |

**Table 2.**
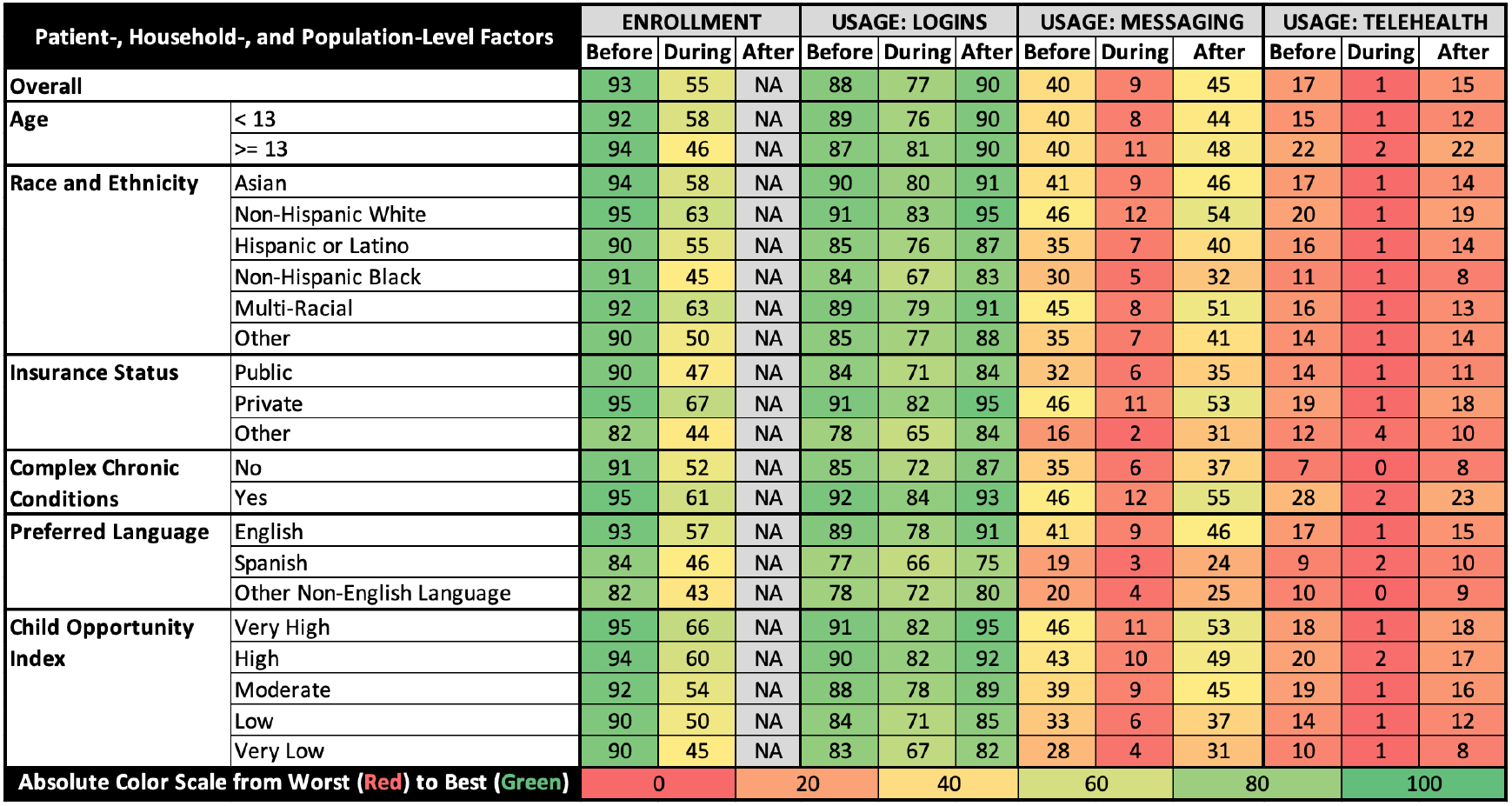
Unadjusted Rates of Portal Enrollment and Usage Before, During, and After Hospitalization (%) “Before” represents enrollment and usage before hospitalization out of the total population. “During” represents enrollment during hospitalization out of those not enrolled prior to admission, and usage during hospitalization out of those enrolled during hospitalization. “After” represents usage after discharge out of those enrolled by discharge. We did not measure enrollment after discharge.

### Enrollment

When examining unadjusted patient portal enrollment before and during hospitalization, we observed lower enrollment rates among patients who identified as Hispanic or Latino or as Non-Hispanic Black, had public insurance, had a household preferred language other than English, or lived in low or very low opportunity neighborhoods. In adjusted multivariable regression models, we found that age <13, identifying as Hispanic or Latino or Non-Hispanic Black race (relative to Asian), public insurance (relative to private insurance), household preferred LOE (relative to English), and high, moderate, low, and very low COI (relative to the highest quintile) were associated with decreased odds of portal enrollment. Presence of one or more CCCs was associated with increased odds of portal enrollment.

Among the subset of patients who were not enrolled in the patient portal prior to admission, we found that age ≥13, public insurance, household preferred LOE, and moderate, low, or very low COI were associated with decreased odds of being enrolled during the hospitalization. (Figure 1)

**Figure 1:**
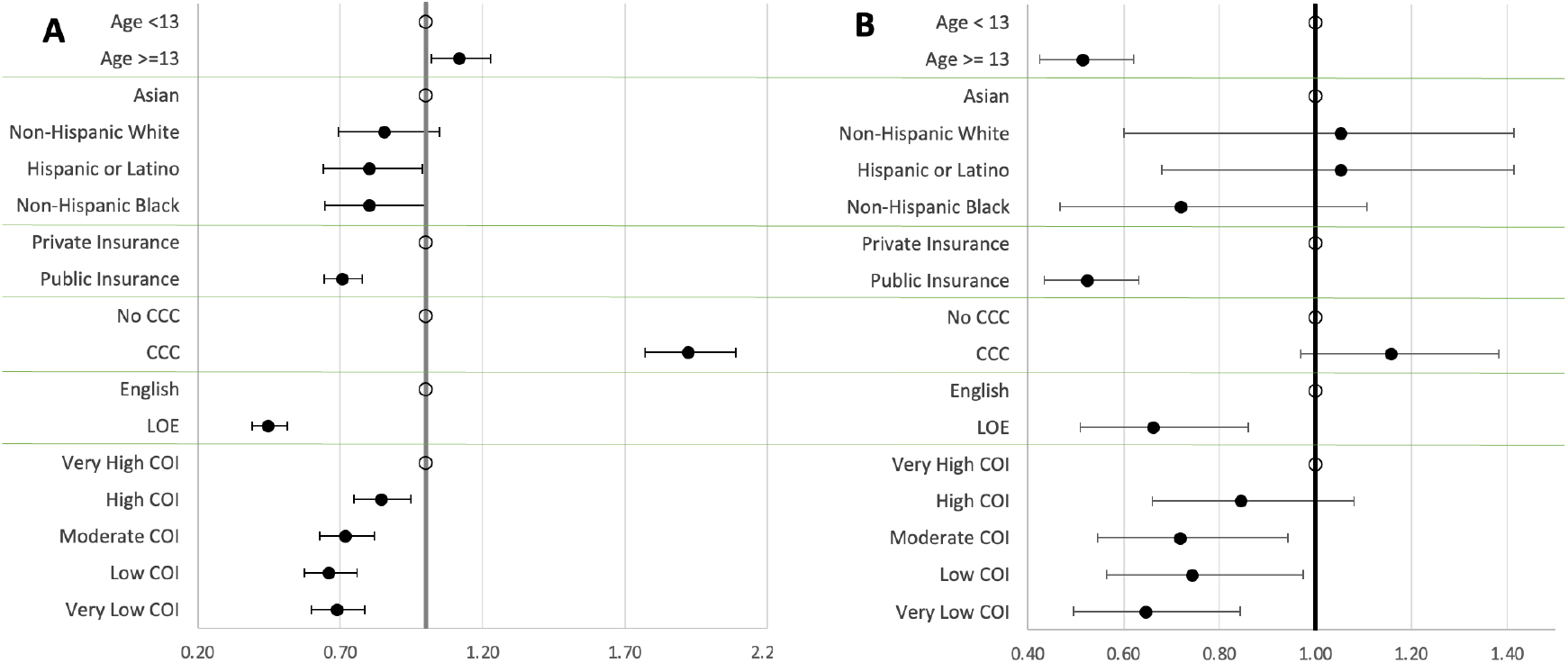
Portal Enrollment Before and During Hospitalization. 1A: Enrollment Before Hospitalization 1B: Enrollment During Hospitalization

### Usage

When examining unadjusted patterns of patient portal utilization, we found relatively lower usage rates among patients who identified as Hispanic, Latino, or Non-Hispanic Black, had public insurance, had a household language other than English, or lived in low or very low opportunity neighborhoods.

In adjusted multivariable regression models assessing usage *before hospitalization* (Figure 2), we found that age ≥ 13 was associated with lower odds of logins and messages before hospitalization, while age < 13 was associated with lower odds of telehealth use. Non-Hispanic Black, Hispanic or Latino, and Non-Hispanic White race or ethnicity (relative to Asian) were associated with lower odds of logins before hospitalization, and Non-Hispanic Black race was associated with lower odds of messaging or telehealth use before hospitalization. Public insurance (relative to private insurance) was associated with lower odds of logins and messages before hospitalization. Household preferred LOE (relative to English), and low or very low COI were both associated with decreased odds of logins, messages and telehealth use before hospitalization. Presence of one or more CCCs was associated with increased odds of patient portal logins, messaging, and telehealth use before hospitalization.

**Figure 2:**
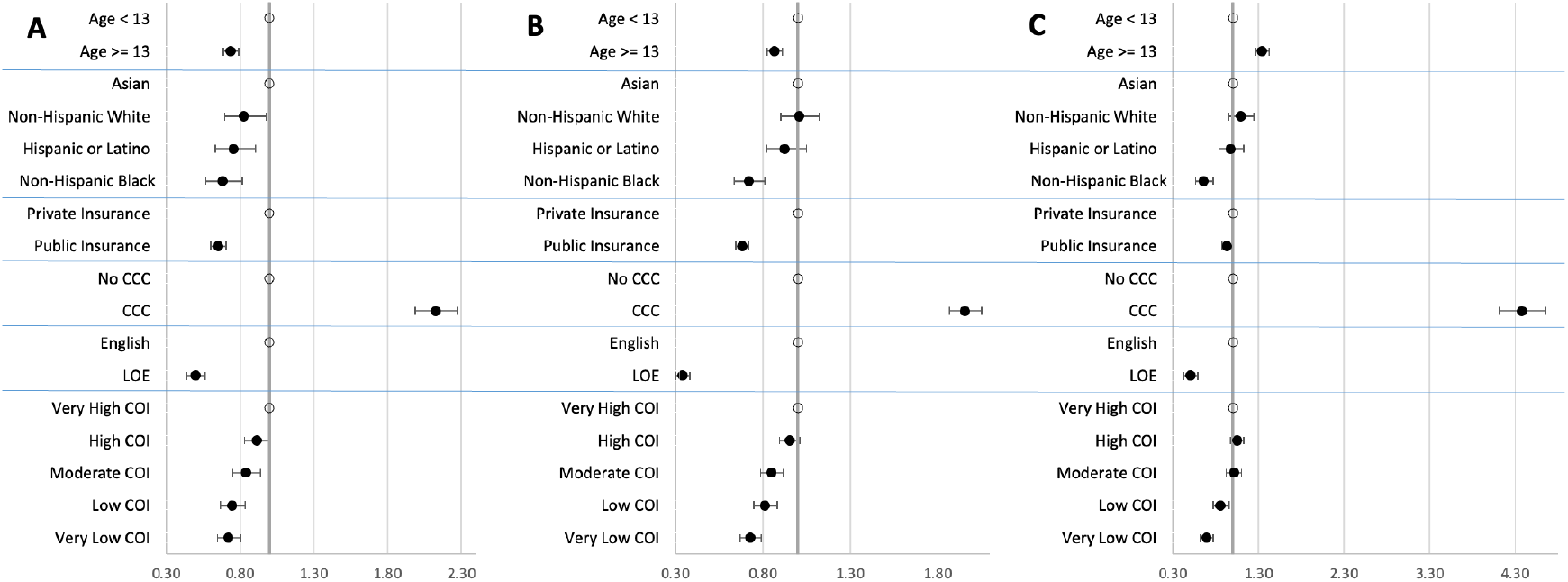
Portal Usage Before Hospitalization. 2A: Logins Before Hospitalization 2B: Messaging Before Hospitalization 2C: Telehealth Before Hospitalization

In adjusted models assessing usage *during hospitalization* (Figure 3), we found that Non-Hispanic Black race was associated with lower odds of logins and messaging during hospitalization. Public insurance, household LOE, and low or very low COI were associated with lower odds of logins and messaging during hospitalization. Presence of one or more CCCs was associated with increased odds of patient portal logins and messaging during hospitalizations. As <2% of enrolled patients utilized telehealth during hospitalization, we excluded this rare event from our analysis.

**Figure 3:**
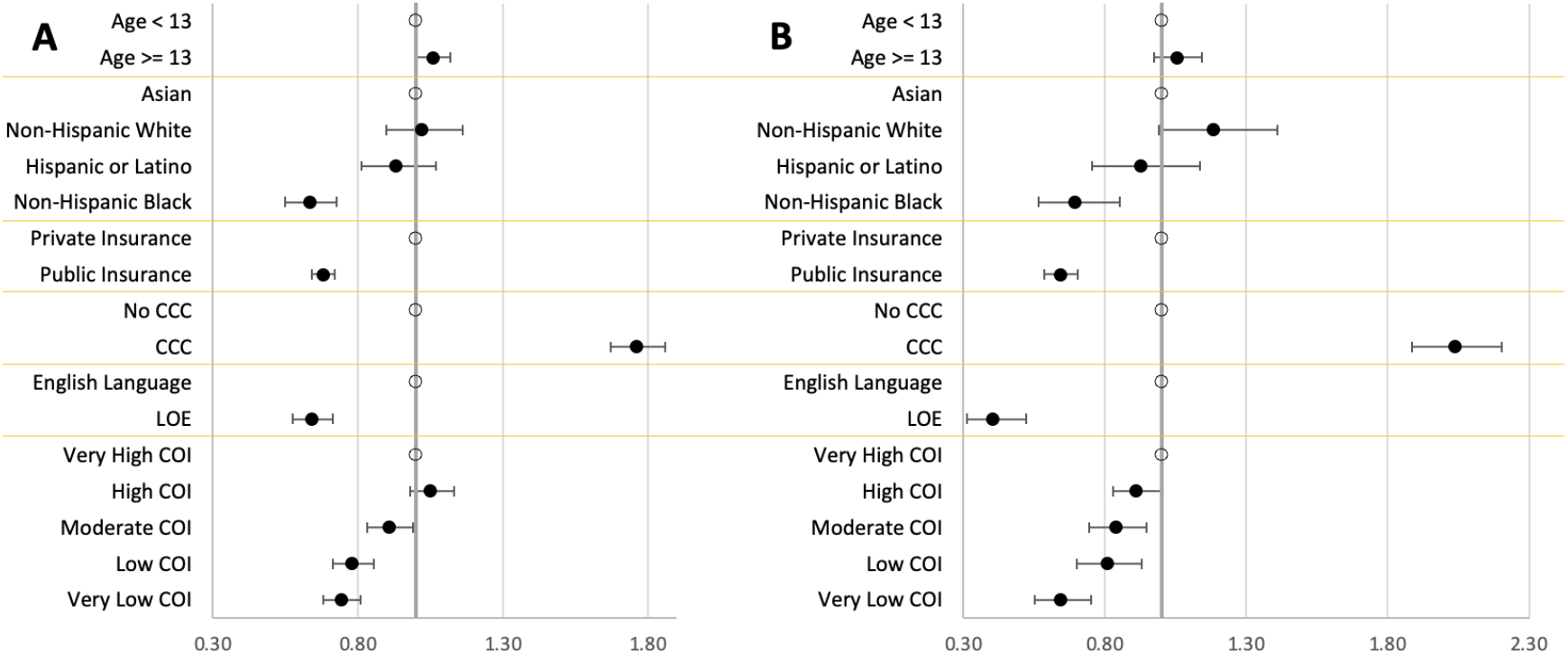
Portal Usage During Hospitalization. 3A: Logins During Hospitalization 3B: Messaging During Hospitalization

In adjusted models assessing usage *after hospitalization* (Figure 4), we found that age ≥ 13 was associated with lower odds of logins, while age < 13 was associated with lower odds of telehealth use. Non-Hispanic White and Non-Hispanic Black race, public insurance, and household preferred LOE were associated with lower odds of logins, messaging, and telehealth after hospitalization. High, moderate, low, and very low COI were associated with lower odds of logins and messaging after hospitalization, while only low or very low COI were associated with lower odds of telehealth after hospitalization. Presence of one or more CCCs was associated with increased odds of patient portal logins, messaging, and telehealth use following hospitalizations.

**Figure 4:**
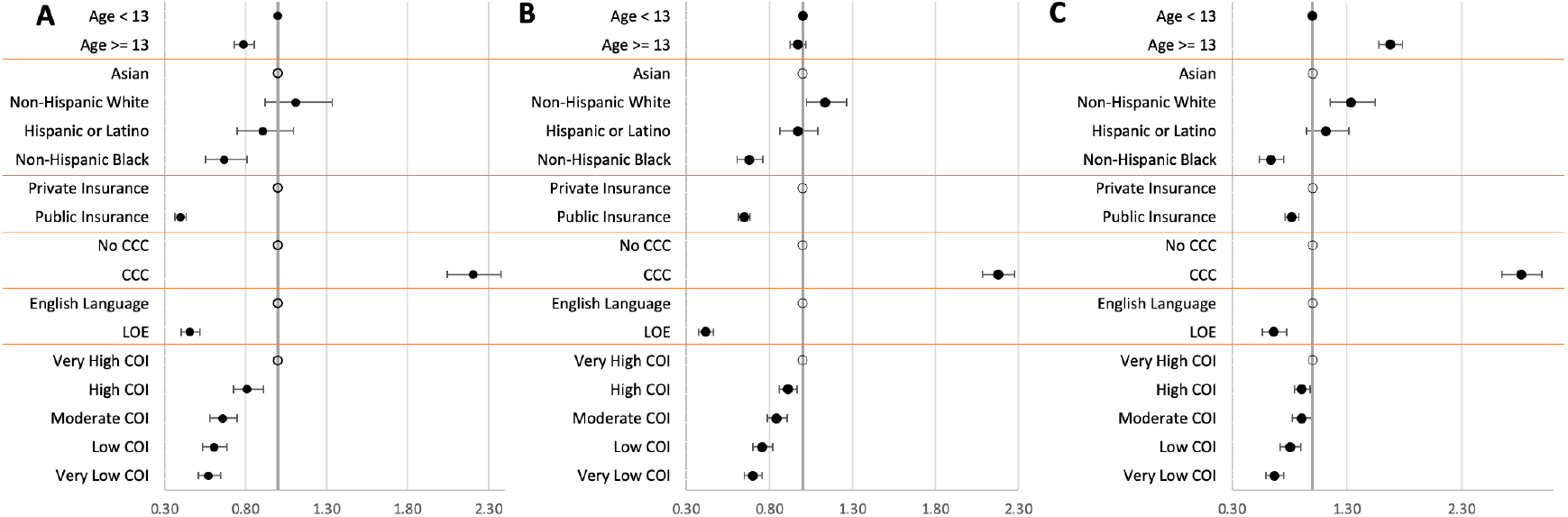
Portal Usage After Hospitalization. 4A: Logins After Hospitalization 4B: Messaging After Hospitalization 4C: Telehealth After Hospitalization

## Discussion

In this observational, cross-sectional analysis of patient portal data for a large, diverse cohort of patients across two children’s hospitals, we found that patients who were publicly insured, had a household preferred language other than English, and lived in lower opportunity neighborhoods were less likely to be enrolled in patient portals both before and during hospitalizations. These groups were also less likely to have portal usage (i.e. logins, sent messages, or telehealth encounters) both before and after hospitalization.

Examining patient-level factors, we noted patients < 13 had lower odds of enrollment before hospitalization, which may reflect fewer prior encounters with the health care system as opportunities for portal invitation or increased portal engagement among teenage patients as compared to caregivers of younger patients. We did find, however, that patients ≥ 13 had lower odds of enrollment during hospitalization, which may suggest the need for more intentional efforts to enroll adolescents. This is particularly important given emerging work highlighting the benefits of technology-based health care for adolescents’ autonomy during the transition into adult care.^21-23^

Despite greater odds of enrollment before hospitalization, users of adolescent accounts were less likely to log in before and after hospitalization, but more likely to engage in telehealth. This may indicate differences in features utilized and prioritized across these two populations. Prior literature has highlighted that parents of younger children were more likely to review appointments, laboratory tests, problem lists, and allergies compared to adolescents themselves,^21^ while there have been increasing opportunities for adolescents to benefit from telehealth.^24^

Prior research has also highlighted decreased patient portal use among racial and ethnic minoritized groups, even when accounting for income and education.^7^ As race and ethnicity are social constructs, these disparities are likely attributable to effects of structural racism not fully captured by adjusting for observable factors such as income and educational level. In our study, we controlled for additional factors including language preference and neighborhood opportunity, which may be mediators of the effects of structural racism on portal engagement. Adjusting for these factors attenuated these observed disparities, particularly for in-hospital enrollment, which may suggest that household LOE and neighborhood COI are reliable metrics to target interventions aimed at narrowing racial and ethnic disparities. Nevertheless, even after adjusting for these additional covariates, patients identifying as Non-Hispanic Black were still less likely to be enrolled before admission and less likely to use their portal before, during, and after hospitalization. This suggests a continued need to better understand upstream digital drivers of health that lead to racial disparities in health technology engagement.^17, 25, 26^

We also found that children with one or more CCCs were significantly more likely to have portal enrollment and usage at all time points. These children often have more frequent appointments and interfaces with the health care system, creating more opportunities for patient portal enrollment or usage.^27^ They may also have a greater perceived value of or need for digital health tools, as telehealth has important benefits for families with children with medical complexity, who often have health care appointments and face disproportionately higher financial burdens for health care-related transportation.^28^

Examining household factors, our findings also suggest that families’ use of LOE was associated with significantly lower portal enrollment and usage. Spanish-speaking families represented the largest cohort of our families using LOE. Our patient portal is available in many languages other than English, including Spanish, but our data show that making portals available in other languages alone is not sufficient to address disparities in patient portal engagement and utilization. Previous studies have shown that Spanish-speaking families experience several barriers to digital health engagement, including decreased awareness of patient portals and other digital health technologies, lack of portal features available in Spanish, and lack of interpreter availability in telehealth encounters.^6^ This should not, however, be interpreted as a lack of interest in portal use, as research in adult populations has shown no significant differences in Spanish-speaking and English-speaking families’ perceptions of the value of these portals when available in their language.^29^ Therefore, improving language accessibility of these platforms, such as the ability to translate provider messages or institution-specific portal resources in preferred languages, and increasing family awareness of such tools are prime areas of opportunity for future quality improvement efforts.

Examining population-level factors, we also found incremental increases in the likelihood of portal enrollment and usage with increasing neighborhood opportunity. This pattern has previously been demonstrated in literature focused on outpatient patient portal engagement among pediatric oncology patients.^30^ Prior research in adult patients has found neighborhood broadband internet access to be associated with portal use.^8^ Our proxy measure of neighborhood opportunity, the COI 3.0, incorporates broadband access as a component metric. Therefore, while we use neighborhood opportunity as a composite estimation of structural drivers of health including access to technology, we cannot reliably differentiate which factors within this index are the most influential. Future research exploring the component domains within the COI may help us better understand the impact of neighborhood opportunity on patient portal use and enrollment. Regardless, the COI appears to be a promising tool to help identify key areas to narrow disparities, perhaps by targeting interventions to improve digital health access and engagement in health care settings serving predominantly lower opportunity neighborhoods. In the inpatient setting, interventions aimed at reducing disparities in patient portal enrollment and use could prioritize individuals from lower COI neighborhoods.

Finally, while we saw similar trends in disparities for both portal enrollment and portal usage, our descriptive results show notable differences between enrollment and usage rates. Understanding these differences is important as there are likely distinct disparities between these processes, which may necessitate distinct improvement strategies. Disparities in enrollment could be attributed to differences including who providers invite to enroll, what clinical settings offer enrollment, or families’ trust in digital health tools. Disparities in usage could be attributed to differences including families’ broadband access or technological or health literacy.

In addition, while not evaluated in this study, there may be substantial differences in the frequency of portal usage among portal enrollees, leading to additional disparities in access to care. While longitudinal engagement with a health system’s portal may not be relevant for all children admitted to a large children’s hospital, we restricted our study population to patients with primary or subspecialty care within our system, in order to identify the subset of patients most likely to benefit from portal enrollment. As portal engagement is a critical step in receiving the intended health benefits of these tools, quality improvement efforts should go beyond efforts to increase enrollment and also aim to increase families’ comfort in utilizing patient portal features to promote meaningful use.

## Limitations

To our knowledge, this is the first study to examine both portal enrollment and portal usage among hospitalized children, who represent a uniquely high-risk population. Several study limitations are worth noting. First, we found that overall rates of portal enrollment and use were higher in our population as compared to prior published studies,^11, 31^ which may reflect differences between health systems’ efforts to promote portal enrollment and could limit the generalizability of our findings. Second, we did not have access to granular data that would allow us to examine usage for all portal features (e.g. viewing labs), distinguish patient or proxy usage, or assess frequency of usage (e.g. number of logins, messages sent, or telehealth visits). Finally, we are unable to link specific aspects of portal use to quality of care metrics or clinical outcomes. Improving our ability to reliably measure clinically beneficial forms of portal use could help us better identify features that quality improvement efforts should focus on in order to reduce disparities in patient portal engagement.

## Conclusions

In this cross-sectional study, we found that although overall rates of patient portal enrollment were high among hospitalized children and their caregivers, public insurance and use of a LOE were associated with lower portal enrollment and usage, while living in high or very high opportunity neighborhoods and having one or more CCCs were associated with greater patient portal enrollment and usage. Improvement efforts aimed at reducing disparities in meaningful use of patient portals should focus on improving access and digital health literacy among families who are publicly insured, use languages other than English, or live in lower opportunity neighborhoods.

## Data Availability

Interested parties can contact the first author for data sharing requests.

